# Time to Develop and Predictors of Peripheral Intravenous Cannula–Related Local Complications Among Neonates Admitted to Neonatal Intensive Care Units in Tigray, Ethiopia: A Prospective Cohort Study

**DOI:** 10.64898/2026.03.20.26348952

**Authors:** Girmay Teklay welesamuel, Hagos Gebreluel, Teklemariam Gebregziabher, Teklewoini Mariye, Guesh Mebrahtom

**Author notes:** Corresponding author (GTW). These authors contributed equally to this work.

## Abstract

**Background:** Peripheral intravenous cannulation is common procedure in neonatal care, yet it carries a significant risk of local complications that can compromise therapy and prolong hospital stay. Understanding the timing and predictors of Peripheral intravenous cannulation related local complications is crucial for improving neonatal outcomes. This study aimed to determine the incidence, timing, and predictors of Peripheral intravenous cannulation related local complications among neonates admitted to public hospitals in the Tigray, northern Ethiopia.

**Methods:** A prospective cohort study was conducted among 528 neonates who underwent peripheral intravenous cannulation. Data were collected using structured questionnaires and observational checklists. Neonates were followed for up to 96 hours. Cox proportional hazards regression was used to identify predictors of local peripheral intravenous cannulation related complications, with Kaplan Meier analysis to estimate complication free survival. Model assumption was assessed using Schoenfeld residuals and goodness of fit evaluated by Cox-Snell residuals, with variables showing p < 0.05 in the multivariable model considered statistically significant.

**Result:** The overall incidence of local peripheral intravenous cannulation -related complications among neonates was 41%, yielding an overall incidence rate of 8.85 per 1,000 catheter-hours. The median time to complication was 78 hours (95% CI: 67–80). The multivariable analysis identified the following independent predictors: chronic illness (AHR=1.54, 95% CI: 1.15–2.07), absence of saline flushing (AHR =1.83, 95% CI: 1.39–2.41), non-visible veins (AHR =2.07, 95% CI: 1.55–2.76), three or more insertion attempts (AHR =1.85, 95% CI: 1.15–2.98), cannula placement in the leg (AHR =1.84, 95% CI: 1.28–2.64), and cubital fossa (AHR =1.62, 95% CI: 1.10–2.39).

**Conclusion:** Local Peripheral intravenous cannulation complications in neonates are common and occur early, particularly among high-risk groups. Intervention such as routine IV-line flushing, careful vein selection, minimizing repeated insertion attempts, and avoiding high risk insertion sites can reduce complications. Close monitoring of neonates with chronic conditions and adherence to cannula replacement guidelines are recommended. Ongoing training for health care providers is essential to improve Peripheral intravenous cannulation care and neonatal outcomes.

## Introduction

Peripheral intravenous cannulation (PIVC) is one of the most commonly performed invasive procedures in clinical practice and play a vital role in the administration of fluids, medications, and blood products (1). It is estimated that billions peripheral intravenous cannula are inserted globally each year, highlighting its importance in modern healthcare delivery(2). In neonatal intensive care units (NICUs), PIVC is particularly essential, as neonates frequently require intravenous therapy for the management of infections, prematurity related conditions, and other critical illness (3).

Despite its widespread use, PIVC associated with arrange of local complications that may compromised treatment effectiveness and patient safety (4). Common complications include phlebitis (2, 5, 6), infiltration (2, 6, 7), extravasation, and occlusion, which can lead to tissue injury, interruption of therapy, repeated cannulation, and prolonged hospitalization (8). Clinical guidelines often recommend routine replacement of cannulas within 72–96 hours to reduce the risk (9).

Neonates are particularly vulnerable to PIVC-related complications due to their fragile and small caliber veins, limited vascular access sites, and physiological immaturity (10). These factors increase the likelihood of cannula failure, necessitating repeated insertion attempts that can cause pain, increase infection risk, and delay essential treatment. The burden of such complications is even more pronounced in resource limited settings, where constrains in skilled personnel. Equipment, and standardized practices may further exacerbate adverse outcomes (4, 11).

Previous studies have reported considerable rates of PIVC complications in pediatric and neonatal populations worldwide(4). Several factors have been identified as contributors to these complications, including insertion site (2, 5, 12, 13), number of cannulation attempts (13, 14), infusion practice (14, 15), vein visibility (16), patient related characteristics(17), and procedural techniques(18, 19). However, most studies in Ethiopia have focused on mixed population of adult and children, limiting their applicability to neonatal care (2, 12, 20). Furthermore, although some neonatal studies report catheter dwell time (4, 21, 22), there is limited use of time to event analysis to better understand the timing and risk dynamics of complications.

This evidence gap is particularly critical in low- and middle-income countries such as Ethiopia, where neonatal morbidity and mortality remain high (23, 24). In the Tigray region, there is scarcity of context specific data on the timing and predictors of PIVC related complications among neonates (25, 26). The absence of such evidence may contribute to delayed recognition of complications, inconsistent clinical practices, and suboptimal neonatal care.

Therefore, generating local evidence on the timing and predictors of PIVC-related complications among neonatal outcomes and guiding clinical decision-making. Understanding when complications occur and identifying modifiable risk factors can support better monitoring strategies, optimize catheter management, and reduce unnecessary repeated procedures. Accordingly, this study aimed to assess the time to develop and predictors of local peripheral intravenous cannula-related local complications among neonates admitted to neonatal intensive care units in public general hospitals of Tigray, northern Ethiopia.

## Methodology

### Study area and period

A hospital based prospective cohort study was conducted to assess the time to development and predictors of local complications related to peripheral intravenous cannula use among neonates admitted to neonatal intensive care unit in public hospitals of Tigray, northern Ethiopia. These hospitals provide neonatal intensive care service including intravenous therapy, oxygen therapy, kangaroo mother care, and management of neonatal infections and prematurity.

### Study population

The source population consisted of all neonates admitted to NICU in public hospitals of Tigray who received peripheral intravenous therapy. The study population included neonates admitted to NICU in the selected hospitals who received peripheral intravenous cannulation and fulfilled the inclusion criteria during the study period.

### Eligibility criteria

All neonates admitted to the NICUs of the selected public general hospitals who required peripheral intravenous therapy and whose parents or guardians provided informed consent were included in the study. Neonates who transferred from other unit or health facilities, neonates with preexisting local PIVC complication and neonates who had PIVCs inserted for temporarily diagnostic procedure were excluded from the study.

### Sample size determination

The sample size was calculated using Stata version 17 based on double population proportion formula, assuming 80% statistical power, 5% margin of error, and a 10% allowance for loss to follow-up. Using the flushing variable from previously published study as the reference (2) the final required sample size was 528 participants (n=528).

### Sampling procedure and sampling technique

From the ten accessible public hospitals in the study area, five hospitals were selected using a simple random sampling technique. The total number of neonatal admissions in these hospitals during February and march 2024 was 1070. The sample size was proportionally allocated to each hospital based on the number of admissions during the two months. within each hospitals individual study participants were selected using a systematic random sampling technique. the sampling interval was determined by total number of neonatal admissions during the study period by the total sample size. Thus, the sampling interval was 2. The study participant was selected randomly from the first two admissions using lottery method. After selecting the first participant, every second eligible neonates admitted to the neonatal unit was included in the study until the allocated sample size for each hospital was achieved.

### Study variables

#### Dependent variable

Time from peripheral intravenous cannula (PIVC) insertion to the development of a local PIVC complication.

#### Independent variable

ϖ Socio demographic characteristics of the neonate and mother includes neonatal age, neonatal sex, maternal age, maternal residence, maternal occupation, maternal educational status, and religion.
ϖ Neonatal clinical characteristics includes Weight of the neonate, gestational age, medical diagnosis of the neonate, disease condition of the neonate, anatomical site of insertion, visibility of the vein and indication of the IV cannula insertion
ϖ Procedural related factors include mode of infusion, flushing/irrigation and PIVC insertion attempt

#### Operational definition

- **Time to PIVC complication**: The time in hours from the insertion of the first PIVC to at least the first observed occurrence of any local PIVC-related complication before 96 hours(2).
- **Event:** The development of any of the PIVC related local complications before 96 hours(2).
- **Censored:** Neonates who did not experience local PIVC related complication during their hospital stay in 96 hours of admission; died, disappeared or left against medical advice before the occurrence of PIVC complication within 96 hours of admission; and who was transferred out before the occurrence of local PIVC complication within 96 hours of admission(2).
- **PIVC complication:** The presence of one or more of the PIVC related local complications (phlebitis, infiltration, extravasation and occlusion) (27, 28).
- **Phlebitis**: The presence of at least two signs: pain, redness, swelling, warmth and palpable cord(27).
- **Infiltration:** The presence of at least two signs: swelling, coolness, slowed infusion, observable leakage of IV fluids from the site (29).
- **Extravasation:** The presence of at least two signs: swelling, coolness, or observable leakage of IV fluids from the site(29).
- **Occlusion:** Inability to infuse fluid or medication through the vein(8).

#### Data Collection Tools and Procedures

Data were collected using an interviewer administered structured questionnaire and observational checklist adapted from previously published studies (2, 12, 30). The structured questionnaire consisted of items assessing socio-demographic characteristics of the neonates and the mothers, clinical and procedural related factors related to the neonates. The observational checklist was used to captured outcome-related variables including the occurrence of phlebitis, infiltration, extravasation and occlusion.

The reliability of the questionnaire items used to determine local PIV Complications was assessed using Cronbach’s alpha, which ranged from 0.75 to 0.86, indicating acceptable internal consistency. Content validity of the data collection tool was ensured through comprehensive literature review and expert evaluation by academic advisors and experienced neonatal nurses.

Study participant was selected using a systematic sampling technique. Starting from the first eligible neonate, every second neonatal admission that fulfilled the inclusion criteria and whose mother provided informed consent was included in the study until the proportionally allocated sample size for each selected hospital was reached.

Each enrolled neonate was followed for a maximum of 96 hours or until the development of a PIVC related complication, whichever occurred first. The type of complication and the time interval (in hours) between cannula insertion and the occurrence of the complication were recorded. Data collection began at the time of the first cannula insertion and continued until the occurrence of the first PIVC related complication.

The PIVC insertion site was assessed hourly for signs of complications using the observational checklist. Data were collected by trained BSc Nursed who were not employed at the study hospital and worked in alternating day and night shifts to ensure continuous observation. Recruitment of eligible neonates took place in the neonatal intensive care units (NICU) immediately upon admission.

#### Data quality assurance

The English version of the questionnaire was translated to Tigrigna and then back translated in to English by language expert to ensure consistency and accuracy. Two supervisors who are MSc nurses were recruited to monitor the data collection process. Training was given for data collectors and supervisors for two days in Adwa town. In addition, supervisors received additional orientation on supervision techniques, consistency checks, and daily data review to ensure completeness and accuracy of collected data.

Before actual data collection, the questionnaire was pre-tested for validity and reliability on 5 % of the calculated sample size (27 neonates) in Adwa general hospital. Based on the result of the pretest, necessary modification was made to improve the clarity and reliability of the tool.

During the data collection period, supervisors checked each completed questioner daily for completeness and consistency. Subsequently, the principal investigator reviewed all questionnaires to ensure clarity and completeness. Finally, the collected data were cleaned and edited before analysis.

#### Data entry and analysis

Data were collected using Kobo data collection tool and exported to STATA version 17 for analysis. The proportional hazards assumption was checked using Schoenfeld residuals, and the overall global test was used to assess the model fitness. Multicollinearity among independent variables was assessed using the Variance Inflation Factor (VIF), which ranged from 1.032 to 2.148, indicating no significant multicollinearity. Both bivariable and multivariable Cox proportional hazards regression analyses were performed to identify predictors of PIVC-related complications. The Kaplan–Meier failure estimator was used to determine the median failure time and the incidence of PIVC local complications was determined. The equality of failure functions between groups was assessed using the log-rank test. Continuous variables were summarized using the median and interquartile range (IQR).

All with a p-value ≤ 0.25 in the bivariable analysis and cross-tabulation counts ≥ 5 in each cell whose Kaplan–Meier curves and log-log plots did not cross between categories, were included in the multivariable Cox regression model to identify independent predictors of PIVC complications. Variables that showed an independent association with the outcome were identified based on adjusted hazard ratios with 95% confidence intervals and a p-value < 0.05. Accordingly, variables with a p-value < 0.05 in the multivariable analysis were considered statistically significant predictors of PIVC complications

#### Ethical consideration

Ethical approval was obtained from the Health Research Review committee (HRERC) of Aksum University College of Health Sciences and specialized hospital with approval number of IRB08/2025 on 24/02/2025. Recruitment of data collector was started directly after getting an official permission letter from Tigray Regional Health bureau on March 03/2025. An approval letter was also obtained from the administration office of the participating hospitals. Written informed consent was obtained from all participants after providing detailed information about the purpose, benefits, and potential risks of the study. Participation was voluntary, and caregivers had the right to withdraw from the study at any time without any consequences.

All collected information was recorded anonymously to maintain confidentiality. Privacy and beneficence were ensured throughout the study, and the data were used solely for research purposes.

## RESULT

### Sociodemographic variables of the neonates and mothers

A total of 528 neonates and their mothers were included in the study, yielding a response rate of 100%. The neonates had a median age of 3 days (IQR: 1–12 days), while the mothers had a median age of 24 years (IQR: 21–28 years). Among the neonates, 312 (59.1%) were male and 130 (24.6%) of male neonates developed the event (local PIVC complication). The majority of mothers, 522 (98.9%), resided in urban areas. Regarding the occupation, (432; 81.8%) mothers were housewives. In terms of religion, 510 mothers (96.6%) were identified as Christian, whereas 18 (3.4%) were Muslim. (See **Table 1**).

**Table 1:** Sociodemographic characteristics of mothers and neonatal patients in the selected public general hospitals of Tigray, Ethiopia, 2024/25 (n = 528)

| Variables | Category | Censored<br>N (%) | Event<br>N (%) | Total<br>N (%) |
| --- | --- | --- | --- | --- |
| Age of the neonates |  | 4(1±12) days | 2(1±11) days | 3(1±12) days |
| Age of the mothers |  | 24(21±29) days | 21(21±28) years | 24(21±28) years |
| Sex of the neonates | Male | 182(34.5%) | 130(24.6%) | 312(59.1%) |
|  | Female | 132(25.0%) | 84(15.9%) | 216(40.9%) |
| Residence of the mother | Urban | 309(58.5%) | 213(40.3%) | 522(98.9%) |
|  | Rural | 5(0.9%) | 1(0.2%) | 6(1.1%) |
| Education level of the mother | Not educated | 183(34.7%) | 125(23.7%) | 308(58.3%) |
|  | Primary school | 63(11.9%) | 45(8.5%) | 108(20.5%) |
|  | Secondary school | 51(9.7%) | 35(9%) | 86(26%) |
|  | College and above | 17(3.2%) | 9(1.7%) | 26(4.9%) |
| Occupation of the mother | House wife | 256(48.5%) | 176(33.3%) | 432(81.8%) |
|  | Governmental | 9(1.7%) | 0(0%) | 9(1.7%) |
|  | Laborer | 4(0.8%) | 0(0%) | 4(0.8%) |
|  | Merchant | 45(8.5%) | 38(7.2%) | 83(15.7%) |
| Religion of the mother | Muslim | 9(1.7%) | 9(1.7%) | 18(3.4%) |
|  | Christian | 305(57.8%) | 205(38.8%) | 510(96.6%) |

### Clinical and procedural related factors

A total of 528 neonates who underwent intravenous cannulation were included in the Cox regression analysis. The median weight of the neonates was 2800 gm (IQR: 2552.5–3000), and the median gestational age was 39 weeks (IQR: 38–40). Regarding the indications for IV cannulation more than half of the neonates 292(55.3%) underwent cannulation for medication administration only. In addition, 184 (34.8%) neonates required both fluid and medication therapy. With respect to the infusion modality, 298 (56.4%) neonates received intermittent infusions. Normal saline was used for flushing IV lines in 347(65.7%) neonates and 104 (19.7%) of them developed the event. Vein visibility during cannulation was reported in 399 (75.6%) neonates among whom 131 (24.8%) of them developed the event. In terms of insertion attempts, healthcare providers required three or more insertion attempts in 238 (45.1%) neonates, of whom 126 (23.9%) developed the event. Regarding the anatomical site of cannula insertion, the most frequently used site was the Dorsum of the hand 190 (36.0%) neonates. (See **Table 2**).

**Table 2:**
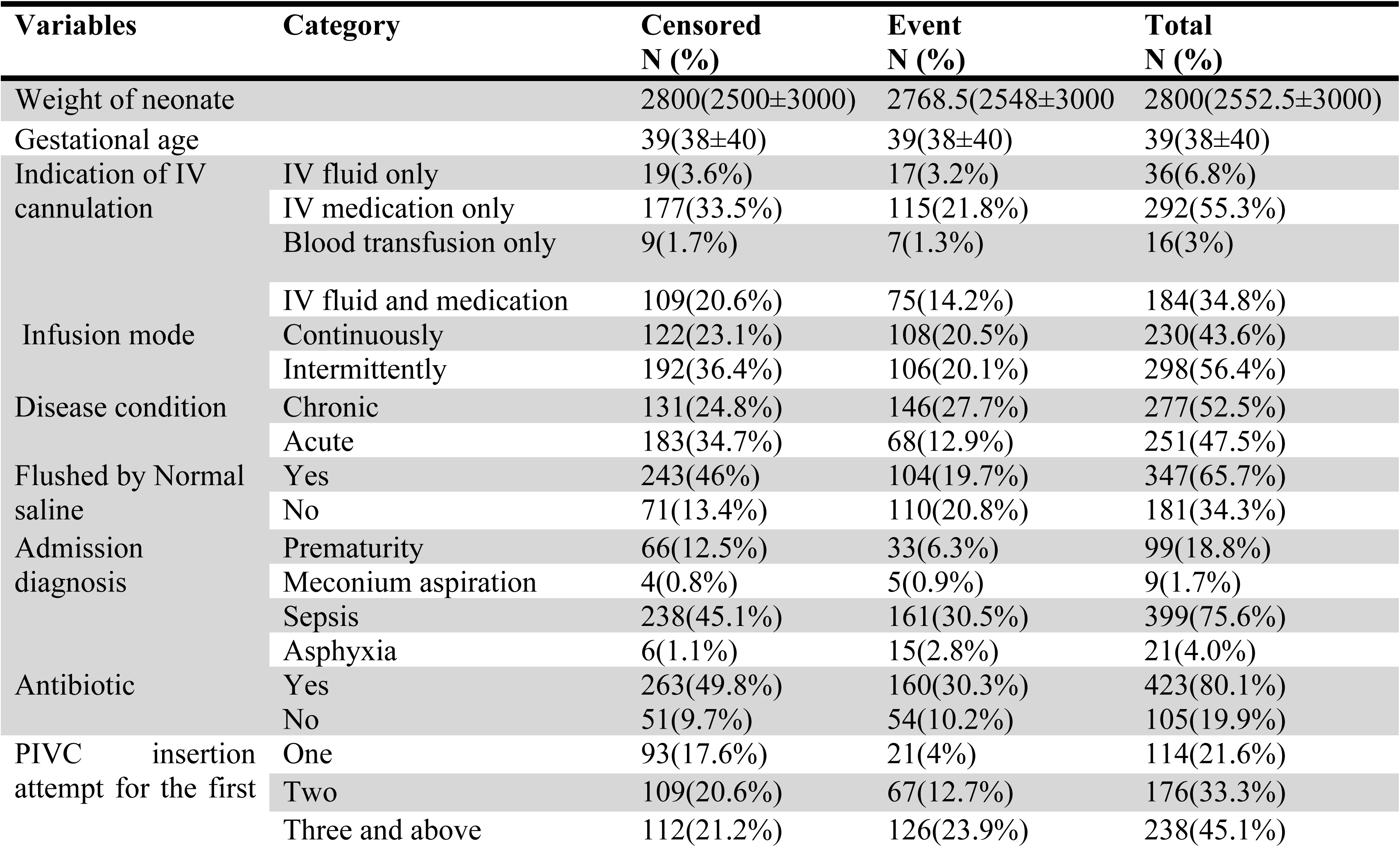
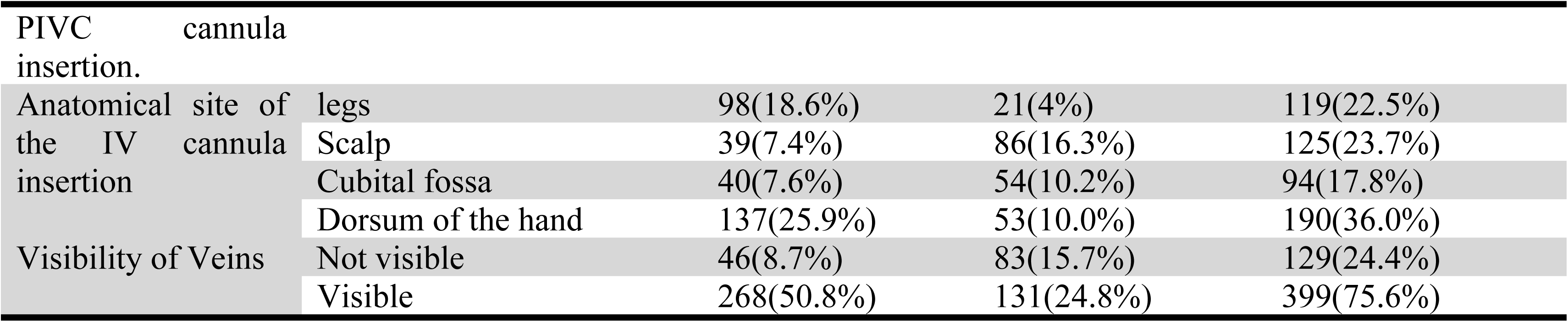
Clinical and procedural related factors of the neonates in selected public general hospitals of Tigray, Ethiopia, 2024/25 (n = 528).

### Incidence rate of the local peripheral IV cannula complication

Among the 528 neonates who underwent intravenous cannulation, 214 (41%) developed local peripheral intravenous cannulation (PIVC) related complications during the follow up period. The duration of follow up ranged from 1 to 96 hours, with the median time to complication of 78 hours (95% CI: 67–80). (Figure 1).

**Figure 1:**
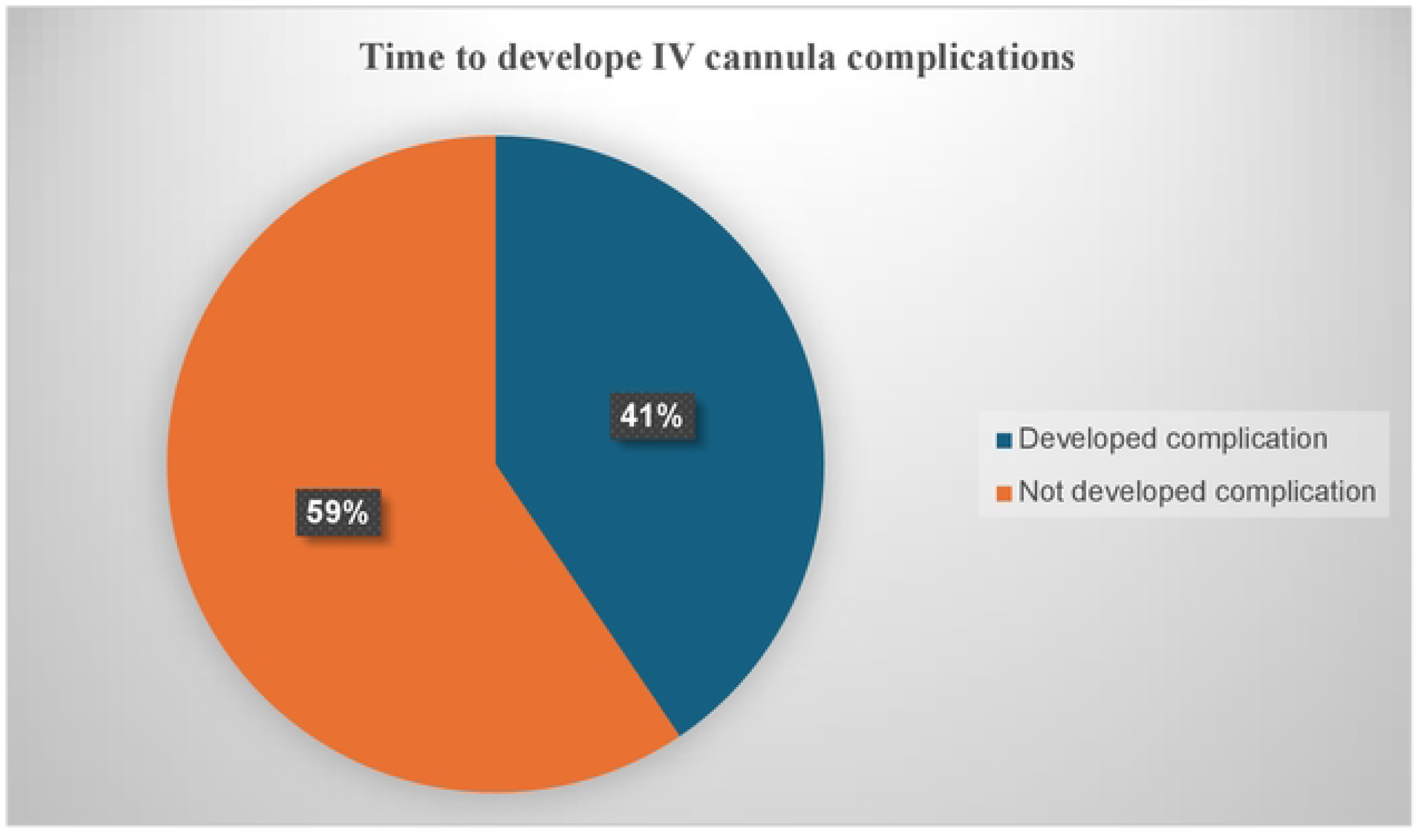
Incidence of local peripheral IV cannula related local complications in neonates admitted in NICUs of public general hospitals of Tigray, Ethiopia

The total person-time at risk contributed by the study participants was 24,212 catheter-hours. Based on this follow up time, the overall incidence rate of PIVC related complications was 8.84 per 1,000 catheter-hours for the first PIVC inserted among the neonates (95% CI: 7.7–10.1). (See ***T*able *3***).

**Table 3:** Incidence rate of local peripheral IV cannula related local complications in neonates admitted in NICUs of public general hospitals of Tigray, 2024/2025.

| Cohort | Person-time | Failures | Rate | 95% Confidence Interval |
| --- | --- | --- | --- | --- |
| Total | 24,212 | 214 | 0.00884 | [0.0077, 0.0101] |

### Log rank test result comparison on different categorical variables

The log-rank test was used to compare the survival experience (time to development of local PIVC complications) among neonates across different categorical variables. The result indicated statistically significant difference in time to develop PIVC related complications among several predictors. Neonates with chronic disease conditions experienced earlier complications compared with those with acute conditions (χ² = 25.69, *p* < 0.001). Similarly, neonates whose IV lines were not flushed with normal saline developed complications earlier than those whose IV lines were flushed (χ² = 38.33, p < 0.001).

A significant difference was also observed according to the number of IV cannula insertion attempts (χ² = 28.40, *p* < 0.001). Neonates who required three or more insertion attempts experienced earlier complications compared with those who required one or two attempts.

Furthermore, the anatomical site of IV cannula insertion showed a statistically significant difference in complication occurrence (χ² = 66.59, *p* < 0.001). In addition, venous visibility during cannulation was significantly associated with the time to development of the complications (χ² = 50.75, *p* < 0.001). (See **Table 4**)

**Table 4:**
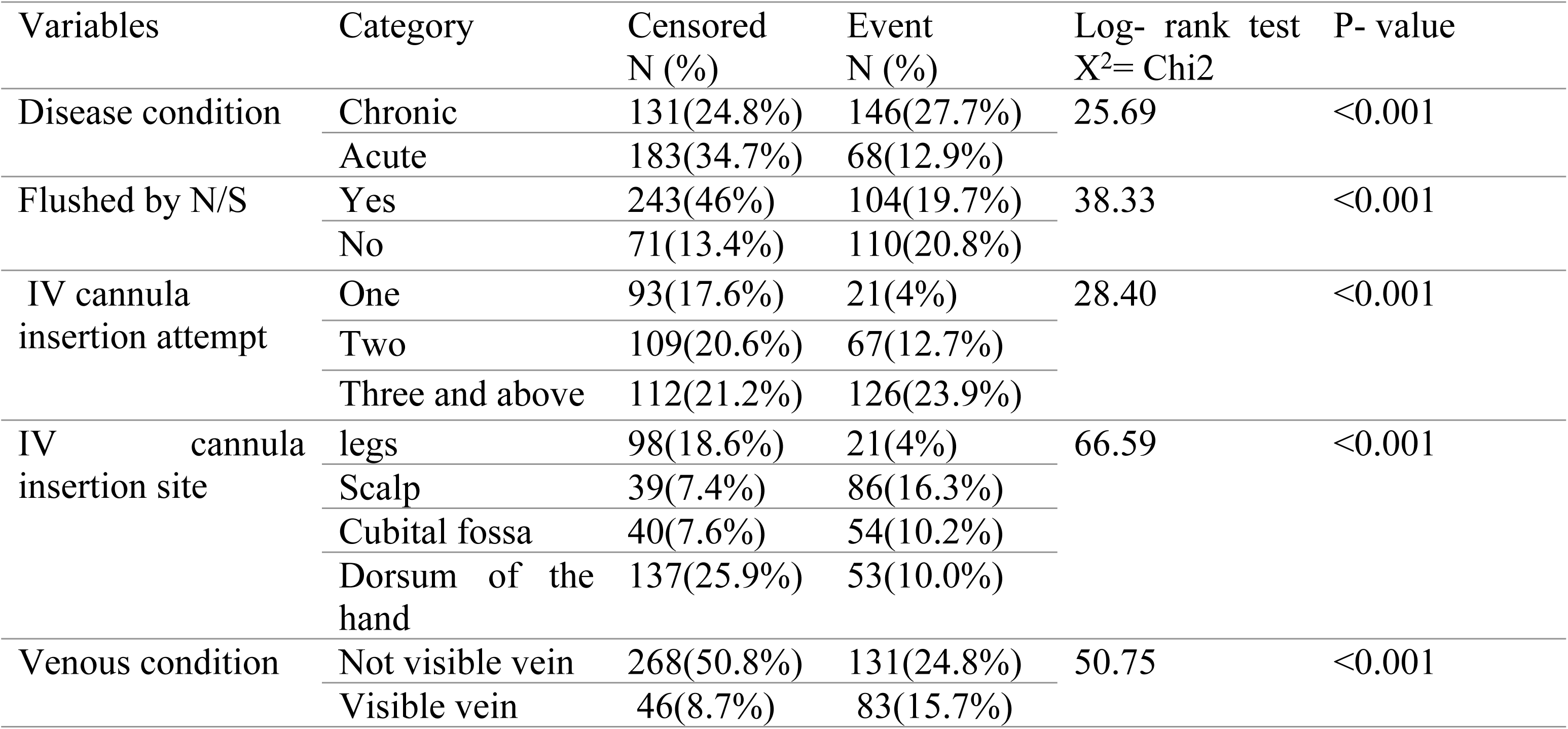
Log rank test result comparison on different categorical variables for local PIVC local complications in selected public general hospitals of Tigray, Ethiopia, 2024/25 (n = 528).

| Variables | Category | Censored<br>N (%) | Event<br>N (%) | Log- rank test<br>X<sup>2</sup>= Chi2 | P- value |
| --- | --- | --- | --- | --- | --- |
| Disease condition | Chronic | 131(24.8%) | 146(27.7%) | 25.69 | <0.001 |
|  | Acute | 183(34.7%) | 68(12.9%) |  |  |
| Flushed by N/S | Yes | 243(46%) | 104(19.7%) | 38.33 | <0.001 |
|  | No | 71(13.4%) | 110(20.8%) |  |  |
| IV cannula<br>insertion attempt | One | 93(17.6%) | 21(4%) | 28.40 | <0.001 |
|  | Two | 109(20.6%) | 67(12.7%) |  |  |
|  | Three and above | 112(21.2%) | 126(23.9%) |  |  |
| IV cannula<br>insertion site | legs | 98(18.6%) | 21(4%) | 66.59 | <0.001 |
|  | Scalp | 39(7.4%) | 86(16.3%) |  |  |
|  | Cubital fossa | 40(7.6%) | 54(10.2%) |  |  |
|  | Dorsum of the<br>hand | 137(25.9%) | 53(10.0%) |  |  |
| Venous condition | Not visible vein | 268(50.8%) | 131(24.8%) | 50.75 | <0.001 |
|  | Visible vein | 46(8.7%) | 83(15.7%) |  |  |

### The cumulative failure probability of PIVC

The cumulative failure probability of Peripheral intravenous cannulation (PIVC) among neonates in public hospitals of Tigray increased steadily over time. Within the first 12 hours, only 20(3.8%) of catheters failed. However, failure probability rises to nearly 50% by 72 hours. By 96 hours, over three-quarters (81%) of catheters had failed. This trend demonstrates a growing risk of PIVC complications with prolonged catheter duration, emphasizing the need for early monitoring and timely replacement (See **Table 5**)

**Table 5:**
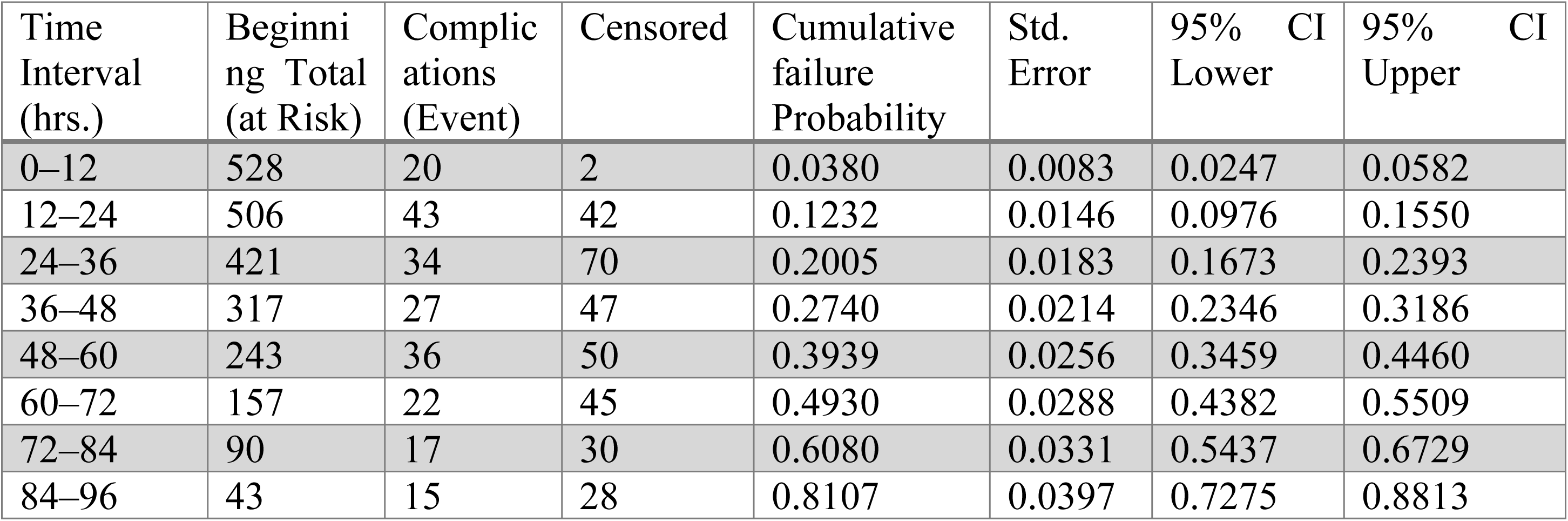
The cumulative failure probability of PIVC among neonates admitted in public hospital of Tigray, Ethiopia in 2024/2025.

**Table 6:** Bivariable and multivariable Cox proportional regression analysis for Time to develop IV cannula complication and its predictor among neonatal patients admitted in selected public general hospitals of Tigray, Ethiopia, 2024/25 (n = 528)

| Variables | Category | Censored N (%) | Event N (%) | CHR (95%) CI | AHR (95% CI) | P-value |
| --- | --- | --- | --- | --- | --- | --- |
| Weight of the neonates |  | 2800(2500 ±3000) | 2768.5(2548 ±3000) | 1.000 (1.00, 1.00) | 1.00 (1.00, 1.00) | 0.389 |
| Infusion mode | Continuously | 122(23.1%) | 108(20.5%) | 1.34 (1.03, 1.76) | 1.27 (0.96, 1.67) | 0.094 |
|  | Intermittently | 192(36.4%) | 106(20.1%) | 1 | 1 |  |
| Disease condition | Chronic | 131(24.8%) | 146(27.7%) | 2.055 (1.54, 2.74) | **1.54 (1.15, 2.07) \*\*** | 0.004 |
|  | Acute | 183(34.7%) | 68(12.9%) | 1 | 1 |  |
| Flushed by N/S. | Yes | 243(46%) | 104(19.7%) | 1 | 1 |  |
|  | No | 71(13.4%) | 110(20.8%) | 2.26(1.73 2.95) | **1.83 (1.39, 2.40) \*\*** | 0.000 |
| Number IV cannula insertion attempt | One | 93(17.6%) | 21(4%) | 1 | 1 |  |
|  | Two | 109(20.6%) | 67(12.7%) | 1.749 (1.07, 2.86) | 1.27 (0.77, 2.10) | 0.343 |
|  | Three and above | 112(21.2%) | 126(23.9%) | 2.89 (1.82, 4.59) | **1.85 (1.15, 2.98) \*\*** | 0.012 |
| IV cannula insertion site | Dorsum hand | 98(18.6%) | 21(4%) | 0.637(0.38 1.06) | 0.61(0.37 0.102) | 0.062 |
|  | Leg | 39(7.4%) | 86(16.3%) | 2.794(1.983, 3.935) | **1.84 (1.28, 2.64) \*\*** | 0.001 |
|  | Cubital fossa | 40(7.6%) | 54(10.2%) | 2.032(1.390, 2.969) | **1.62 (1.10, 2.39) \*\*** | 0.014 |
|  | Scalp | 137(25.9%) | 53(10.0%) | 1 | 1 |  |
| Venous visibility | Not visible | 268(50.8%) | 131(24.8%) | 2.607 (1.98, 3.44) | **2.07 (1.55, 2.76) \*\*** | **0.000** |
|  | Visible |  |  | 1 | 1 |  |

The Kaplan–Meier failure estimate graph shows the cumulative probability of IV cannula complications occurring over the analysis time, which ranges from 0 to 96 hours. The blue curve steadily rises, indicating an increasing risk of complications as time progresses. The failure probability starts near zero and reaches approximately 0.81 by the end of the observation period, reflecting that about 81% of neonates experienced complications by 96 hours. This curve highlights the time-dependent nature of IV cannula complications in neonates. (**See Figure 2**)

**Figure 2:**
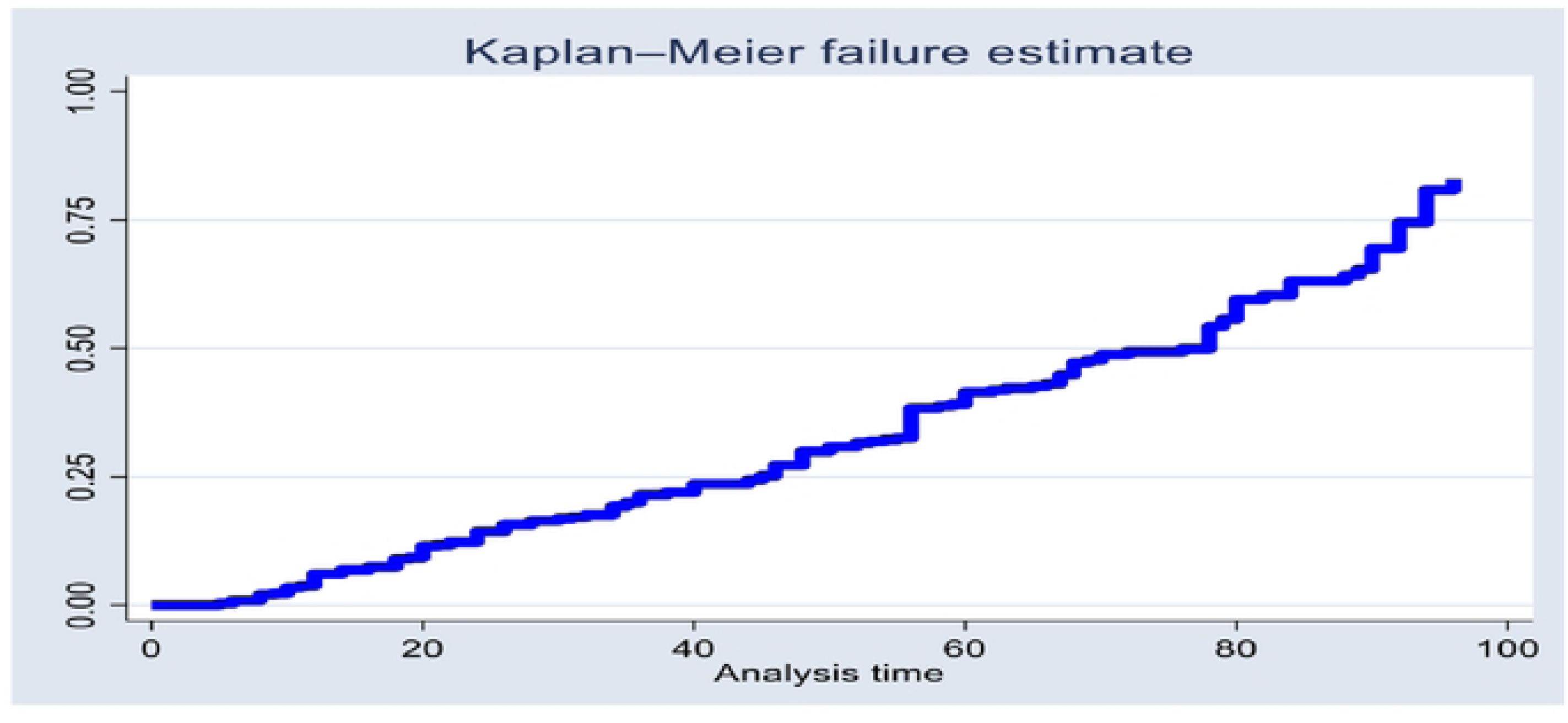
Overall Kaplan-Meier failure estimate of neonatal patients showing Time to develop local IV cannula complications in selected public general hospitals of Tigray, Ethiopia

The red curve (three or more attempts) rises the fastest, indicating that neonates who required three or more IV attempts experienced complications earlier and with a higher cumulative probability over time compared to the other groups. The green curve (two attempts) shows an intermediate risk, whereas the blue curve (one attempt) rises the slowest, suggesting that neonates with a single IV attempt had the lowest and latest incidence of complications. Overall, this graph demonstrates that an increasing number of IV cannulation attempts is associated with earlier and greater risk of IV complications in neonates. (**See Figure 3**)

**Figure 3:**
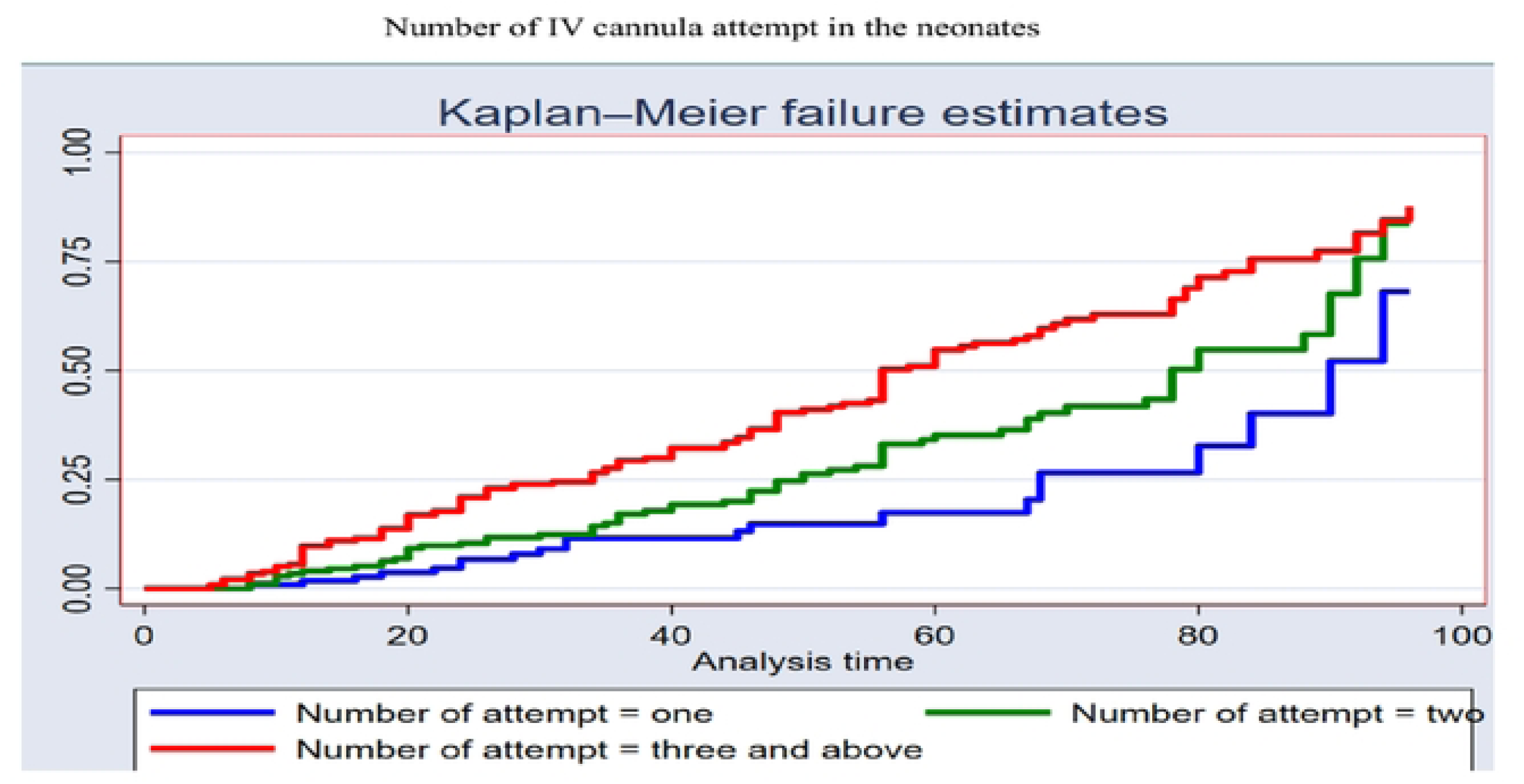
Kaplan–Meier failure estimates depicting the time to develop IV complications based on the number of IV cannula attempts among neonatal patients in selected public hospitals of Tigray, Ethiopia

This Kaplan–Meier failure estimate shows that individuals with non-visible veins (red line) experience failures earlier and more frequently than those with visible veins (blue line). The red curve rises faster, indicating a higher cumulative failure probability over time. By the end of the analysis period, failure probability is around 90% for non-visible veins vs. 70% for visible veins, suggesting that non-visible veins are associated with a higher risk to develop complications. (See **Figure 4**)

**Figure 4:**
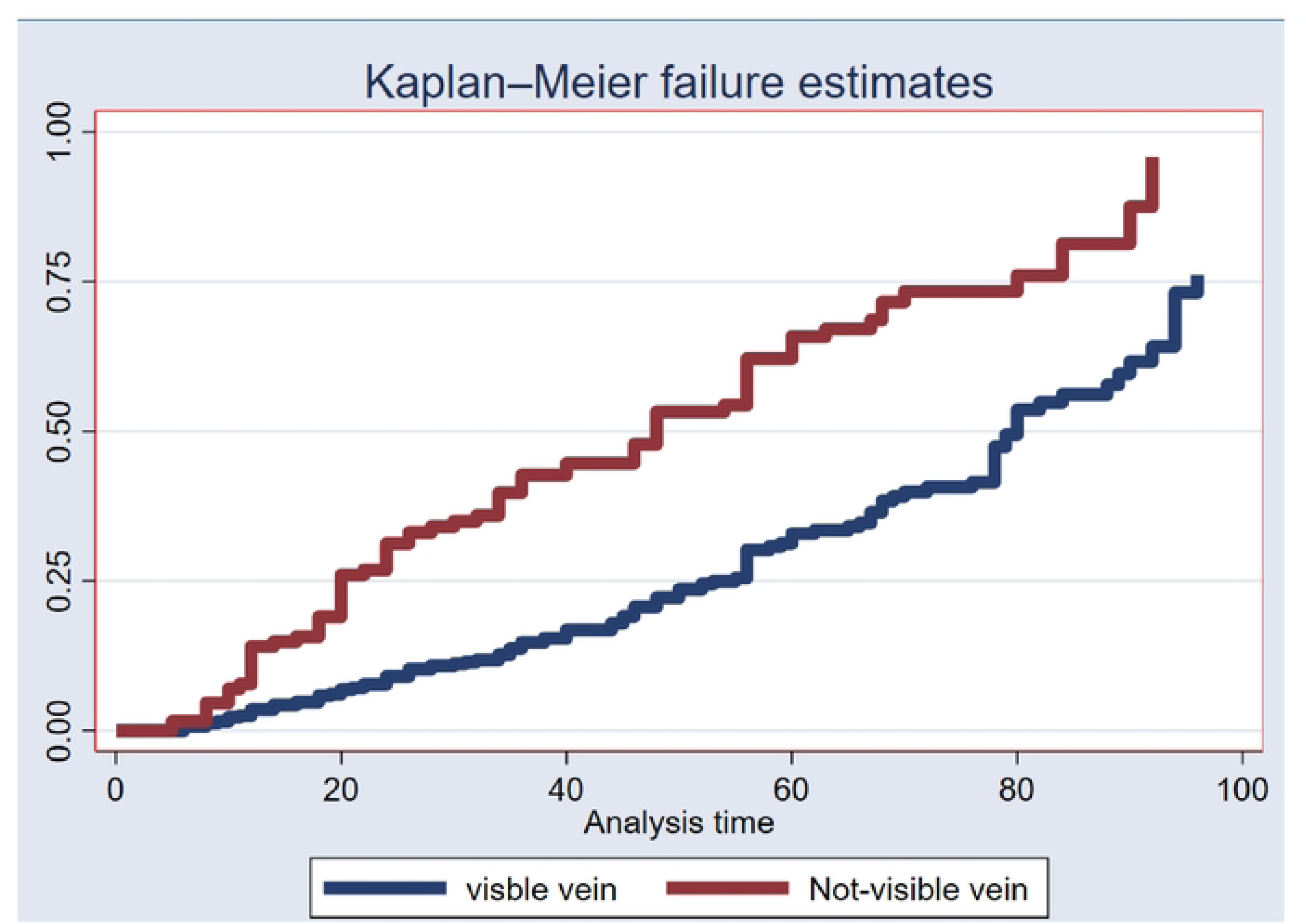
Kaplan-Meier failure estimates of time to develop IV complications by vein-Visibility among neonatal patients in selected public of hospitals of Tigray, Ethiopia

The Kaplan–Meier failure estimate graph compares IV failure rates over time based on different insertion sites. The leg site (red line) shows the highest and earliest failure rate, followed by the cubital fossa (green), scalp (orange), and dorsum of the hand (blue), which has the lowest failure rate. This suggests that IV insertions in the leg are most prone to failure, while those on the hand are the most reliable over time. (See **Figure 5**)

**Figure 5:**
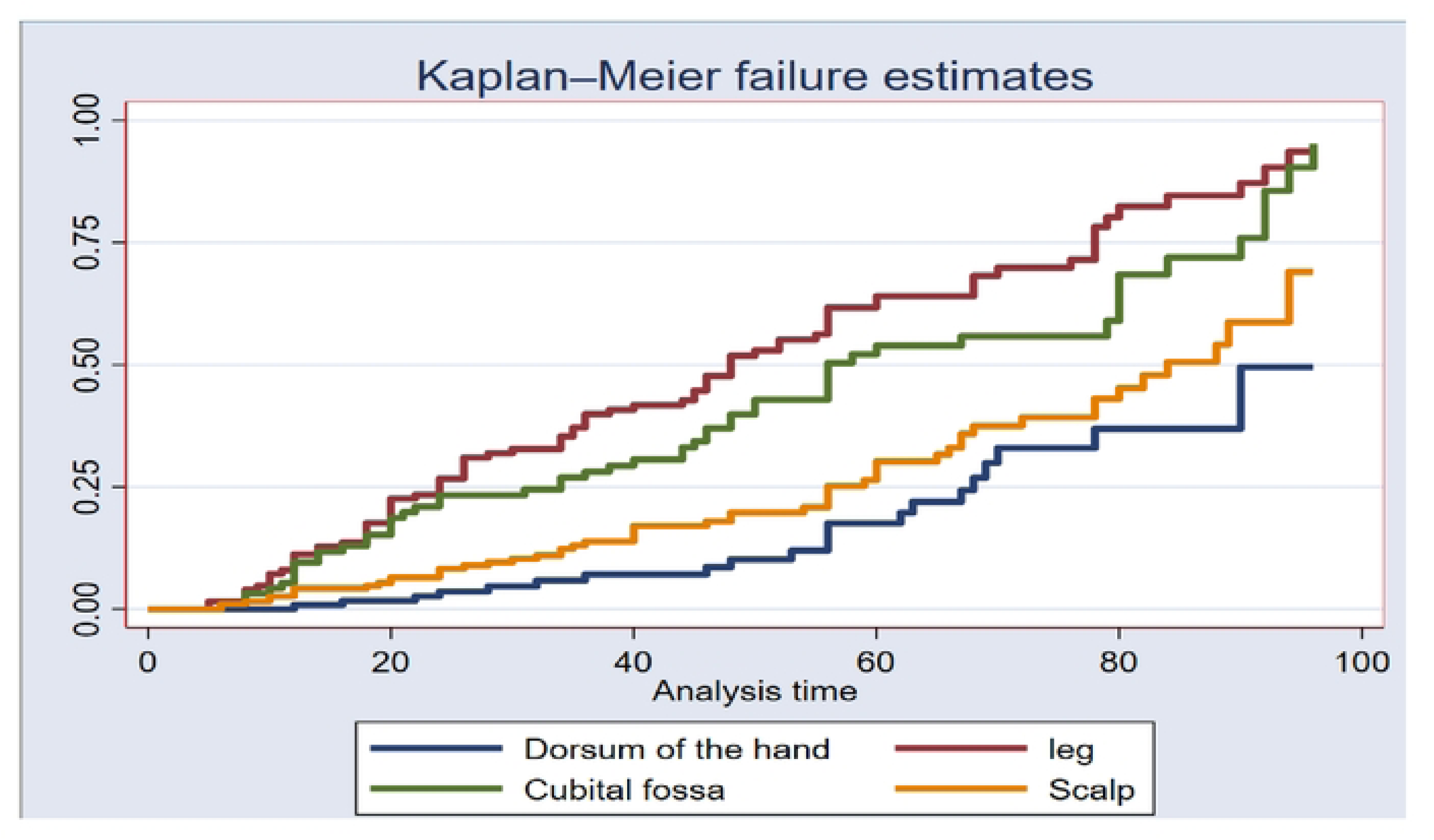
Kaplan-Meier failure estimates for Time to develop of IV complications by insertion site among neonatal patients in selected public hospitals of Tigray, Ethiopia

The graph shows that individuals with non-flushed IVs (blue line) experience failures earlier and more frequently over time than those with flushed IVs (red line). By the end of the analysis, the failure probability is higher for the non-flushed IV group, suggesting that not flushing the IV is associated with an increased risk of complications. (See **Figure 6**)

**Figure 6:**
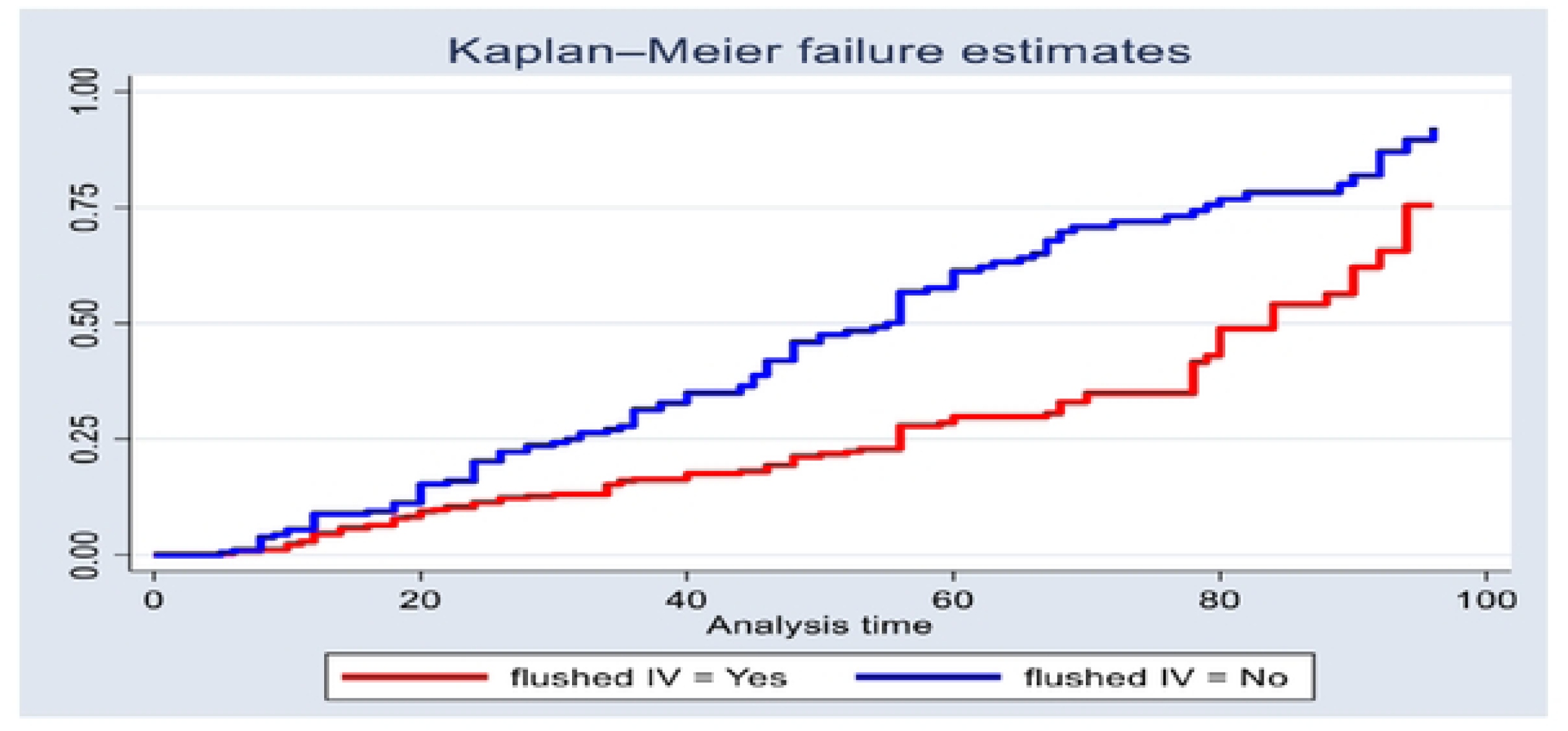
Kaplan-Meier failure estimates comparing time to develop IV cannula complication rates between flushed and not-flushed IV Lines among neonates in selected public hospitals of Tigray, Ethiopia

The Kaplan-Meier curve shows neonates with chronic illnesses experience rapidly increasing complication risks early in treatment, with the failure probability rising sharply within the first time period before stabilizing at a slower rate. This highlights their vulnerability to early complications, underscoring the need for close initial monitoring and prompt intervention. (See **Figure 7**)

**Figure 7:**
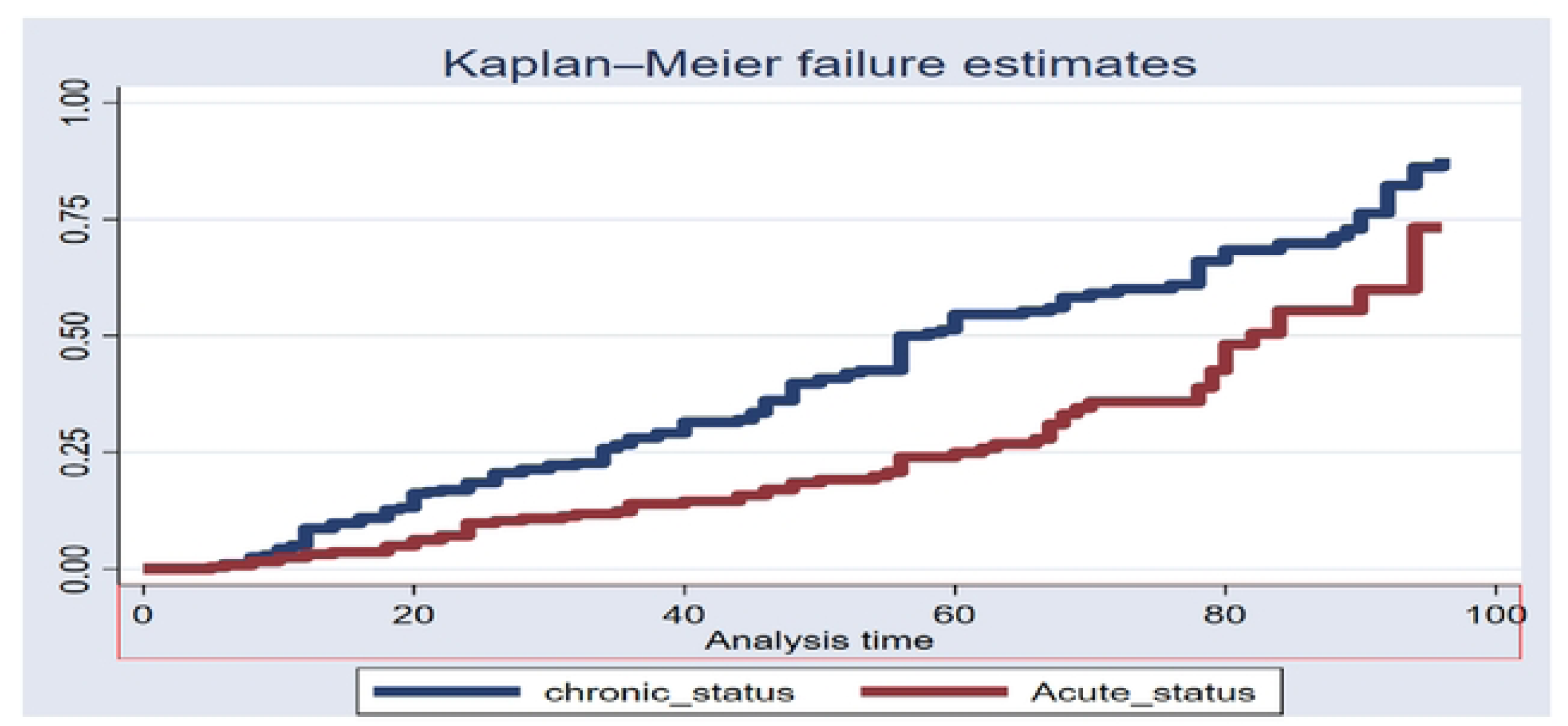
Individual Kaplan-Meier failure estimates for Time to develop of IV cannula complications by disease condition among neonatal patients in selected public general hospitals of Tigray, Ethiopia

### Testing the Model Goodness of Fitness

The graph displays the Cox-Snell residuals plotted against the cumulative hazard function (H) to assess the goodness-of-fit of the Cox proportional hazards model. The close alignment of the Cox-Snell residual curve with the 45° reference line indicates that the model fits the data well, as the observed residuals follow the expected pattern under a correctly specified model. This suggests that the proportional hazards assumption is appropriate and that the model adequately captures the underlying relationships in the data. The strong agreement between the residuals and the reference line supports the validity of the model’s predictions and its suitability for analyzing the time-to-event data in this study (see **Figure 8**).

**Figure 8:**
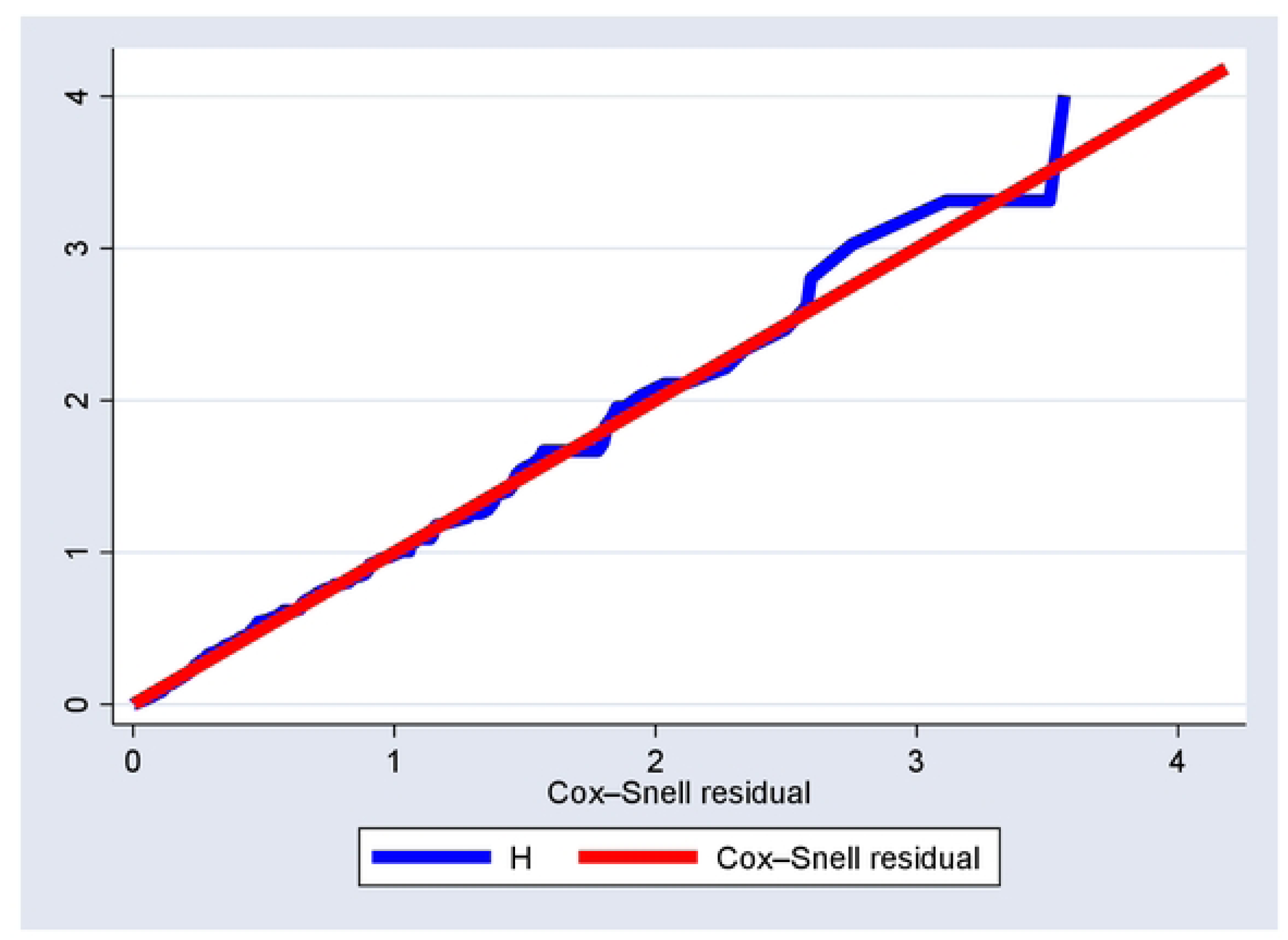
Cox-Snell residual graph for the goodness of model fitness in selected public general hospitals of Tigray, Ethiopia

### Test of Proportional Hazard Assumption

The proportional hazards assumption was formally tested using Schoenfeld residuals test. The overall global test showed no significant violation of the assumption (p-value = 0.556) confirming that the Cox model is valid for analyzing time to event data in this study.

### Predictors of IV cannula complications

The multivariable Cox proportional hazards analysis identified several significant predictors of intravenous cannula (PIVC) complications among neonatal patients. Neonates with chronic illnesses were significantly more likely to develop PIVC complications than those with acute conditions, with a 54% higher risk (AHR = 1.541, 95% CI: 1.147–2.070). Neonates whose IV lines were not flushed had an 82% increased risk of complications (AHR = 1.825, 95% CI: 1.386–2.405) than those flushed. The likelihood of IV complications was more than doubled (AHR = 2.072, 95% CI: 1.552–2.764). IV cannula insertion attempt of three and above was found to increase the risk of developing local PIVC complications by 85% (AHR=1.848 CI: 1.147, 2.979) compared to only one attempt.

## DISCUSSION

Peripheral intravenous cannulation (PIVC) is one of the most common procedures in neonatal care, yet it carries a substantial risk of local complications that can compromise treatment and prolong hospitalization. In this study, we found that PIVC complications among neonates in Tigray, northern Ethiopia occurred early and frequently, with nearly half of the catheters failing with in the first four days after insertion. The overall incidence rate of local PIVC complications was 214(41%), corresponding to 8.85 per 1,000 catheter-hour observation. This finding has important clinical implications, particularly in resource-limited settings such as Ethiopia. A high rate of PIVC complications may lead to repeated cannulation attempts, interruption of essential intravenous therapy, increased pain and stress for neonates, and prolonged hospital stays. Furthermore, resource-constrained healthcare facilities often face shortages of trained staff, limited availability of vein-visualization technologies, and inconsistent adherence to standardized catheter care protocols, all of which may contribute to higher complication rates.

The median time to complication was 78 hours (95% CI: 67–80). These findings underscore the time sensitive nature of catheter-related complications in neonates and highlights the critical need for vigilant monitoring, evidence-based insertion practice, and timely interventions to improve catheter survival and neonatal outcomes. This median survival time higher than previously reported studies in Ethiopia, including 46 hours in both Debre tabor comprehensive specialized hospital and in Bahirdar, and 48 hours in Gurage zone (2, 12, 31). The discrepancy may be due to difference in sample size variation, methodology and recent advancements in neonatal nursing care. The presence of specialized neonatal nurses may have contributed to prolonged catheter survival times by ensuring proper insertion and maintenance practices.

Flushing of IV line with normal saline emerged as a significant predictor of PIVC complications. Routine flushing of IV lines with normal saline is strongly recommended. In this study, neonates whose IV line were not flushed had an 83% higher hazard of complications compared with those whose IV line were flushed (AHR = 1.83, 95% CI: 1.39–2.40). This finding is consistent with a prospective study conducted in Debre tabor comprehensive specialized referral hospital, Northwest Ethiopia (2). Salin flushing likely reduce complication by preventing clot formation, fibrin sheath development, and mechanical irritation of the vein, thereby maintaining catheter potency and reduce local complications(32). These findings underscore the importance of incorporating routine flushing into standard neonatal IV care protocol to improve catheter survival and patient outcomes.

Disease condition played a notable role in neonatal PIVC complication. Neonates with chronic illnesses had a 54% higher hazard of complications compared with those with acute conditions (AHR = 1.54, 95% CI: 1.15–2.07). This aligns with study conducted in Ethiopia, Quatar and Turkey, where chronic illness prolonged intravenous therapy, and complications (33–35). Neonates with chronic conditions are physiologically unstable and immunocompromised, increasing their vulnerability to phlebitis, infiltration, and other local complications (36, 37).

The anatomical site of cannula insertion was another important predictor. Clinicians should prioritize the dorsum of the hand or the scalp for peripheral intravenous catheter (PIVC) insertion, as these sites were found to be the most reliable in minimizing complications. Conversely, insertion in the lower extremities and the cubital fossa should be avoided, when possible, given that catheters placed in the leg were associated with 84% (AHR = 1.84, 95% CI: 1.28–2.64) and 62% (AHR = 1.623, 95% CI: 1.101–2.391) higher hazards respectively as compared with scalp insertion. These findings are consistent with studies conducted in Northwest Ethiopia, Jerusalem, Qatar, Nepal and China(5, 19, 38–40), which report increased risk may be due to anatomical and physiological factors in which the cubital fossa is near flexion joints, where repeated limb movement can cause vein collapse or infiltration, while leg veins are smaller, deeper, and more prone to movement, increasing the risk of thrombophlebitis, occlusion or dislodgement (17, 41). These findings underscore the importance of site selection in improving catheter longevity and reducing patient risk.

Venous visibility during insertion was strongly associated with PIVC complications. Neonates with non-visible veins had more than twice the hazard of complications compared with those with visible vein (AHR=2.07, 95% CI: 1.55–2.76). This result is in line with study conducted in Bahir Dar City, Northwest Ethiopia (12). The association between non visible veins and PIVC complications may be explained by endothelial injury during difficult cannulation. When veins are poorly visible, health care providers are more likely to perform repeated punctures or manipulate the catheter, which can damage the vascular endothelium and trigger inflammation, increasing the risk of phlebitis, infiltration, and extravasation. Neonatal veins are particularly fragile, making them more susceptible to such injury. Similarly, multiple insertion attempts may cause mechanical trauma to the vein and surrounding tissues, leading to endothelial disruption, thrombosis, and catheter instability. These mechanisms may explain the higher risk of complications observed among neonates requiring repeated cannulation attempts (42, 43). Use of vein transillumination and proper training may reduce these risks.

Effort should be made to achieve successful cannulation on the first or second attempt, as multiple insertion attempts significantly increase the risk of complications. Our finding indicated that neonates who required three or more IV cannula insertion attempts had an 85% higher hazard of developing complications compared with those requiring only one attempt (AHR=1.85 CI: 1.15, 2.98). Similar associations have been reported in Bahirdar city, North west Ethiopia (12). This elevated risk is likely due to endothelial injury and mechanical trauma caused by repeated punctures. These results highlight the importance of skilled insertion techniques and appropriate vein selection to minimize the need for repeated attempts.

## CONCLUSION

Local complications related to Peripheral intravenous cannula in neonates occurred at a substantial rate, with a median time to complication of 78 hours. This highlights a relatively high and early risk of catheter failure, supporting for routine PIVC replacement within 72-96 hours. Chronic illness, absence of saline flushing, multiple cannulation attempts, cannula placement a and poor venous visibility were significant predictors of complications.

Continuous education and skill development for neonatal IV insertion and maintenance are recommended. Future studies should consider multicenter and longitudinal designs to evaluate long term outcomes and validate predictors of neonatal PIVC complications. These efforts are critical to improving catheter survival, patient safety, and overall quality of neonatal care. Use of vein-visualization technologies, such as transillumination or near-infrared devices, is strongly recommended to improve first-attempt success and reduce mechanical trauma.

### Limitations of the study

This study was limited by a follow-up period of 96 hours, which may not capture longer-term complications or delayed PIVC related complications. in addition, although data collectors were trained nurses, assessment of local PIVC complication was relied on observational checklist, which could introduce observer subjectivity and potential bias.

## Declarations

### Ethical approval and consent to participate

Ethical approval was obtained from the Health Research Ethical Review Committee (HRERC) of the Aksum University College of Health Sciences and comprehensive Specialized hospital (CHS-SRH), Ethiopia (approval number IRB08/2025). Approval was secured before the initiation of data collection. After that an official support letter was secured from Tigray Regional Health Bureau. Written informed consent was obtained from all participants prior to their inclusion in the study. Participant was voluntary, and confidentiality of the participants information was maintained throughout the study.

### Patent Consent for publication

Not applicable

### Availability of data and materials

All data relevant to the study are included in the article or uploaded as supplementary information

## Data Availability

All relevant data are within the manuscript and its Supporting Information files.

## Acknowledgment

The authors would like to thanks Aksum University college of health science, the Tigray regional health Bureau, and the participating hospitals for their support in facilitating this research. We extend our sincere gratitude to the data collectors, supervisors, and all study participants for their cooperation and valuable contribution

## Authors’ contributions

As the study’s lead investigator, GTW and HG Conceptualization, Data curation, Formal analysis, Investigation, Methodology, Project administration, Resources, Software, Supervision, Validation Visualization, Writing – original draft, Writing – review & editing. TM, GM, and TG Data curation, Formal analysis, Investigation, Methodology, Software, Supervision, Validation, Visualization

